# Performance of cellular senescence measure, p16, and DNA methylation clocks in a clinically relevant model of age acceleration

**DOI:** 10.1101/2023.05.06.23289535

**Authors:** Mina S. Sedrak, Anne Knecht, Susan L. Strum, Canlan Sun, Yuan Chun Ding, Jingran Ji, Thomas A. White, Kirsten Nyrop, Nathan K. LeBrasseur, Susan L. Neuhausen, Natalia Mitin, Hyman Muss

## Abstract

Cellular senescence and DNA methylation are primary aging mechanisms emerging as a potential means of monitoring human aging and evaluating interventions thought to either accelerate or slow an individual’s aging trajectory. However, it is largely unknown whether cellular senescence and signatures of methylation of the specific CpG islands that comprise various epigenetic clocks correlate in humans. We have measured the cellular senescence biomarker, p16 and the five most used epigenetic aging clocks in 251 patients with breast cancer, 49 age-matched non-cancer controls, and 48 patients undergoing cytotoxic chemotherapy treatment. Chemotherapy, a known clinically-relevant inducer of aging, increased expression of p16 but not levels of the most common epigenetic clocks (DNAm-Horvath, PhenoAge, GrimAge, mPoA), with the exception of DNAm-Hannum. Chemotherapy-induced changes in p16 were associated with increased levels of a subset of SASPs, PARC, TNFRII, ICAM1, and TNFa. Cross-sectionally, there was weak to no correlation between p16 expression and epigenetic clocks in cancer patients or non-cancer controls. GrimAge and PhenoAge were the most correlated with p16 (r<0.3), with no correlation between p16 and the pace of aging epigenetic clock. Together, these data show that there is a general discordance between measures of cellular senescence and epigenetic clocks with the senescence marker p16 but not epigenetic clocks of aging responding to a clinically relevant inducer of human aging, cytotoxic chemotherapy.

## INTRODUCTION

Biological mechanisms of aging underlie the appearance of functional decline and chronic age-related conditions. Markers of cellular senescence, components of the senescence-associated secretory phenotype (SASP), and epigenetic clocks based on DNA methylation have emerged as measurements of aging in humans^1^.

Cellular senescence plays an essential role in aging as both an indicator and cause of aging-related functional decline^2–5^. Induced by stimuli such as mitochondrial dysfunction, telomere shortening, hypoxia, disrupted autophagy, and nutrient deprivation, senescent cells undergo irreversible growth arrest but remain metabolically active, secreting soluble factors that promote inflammation and fibrosis and further induce senescence in healthy cells both locally and at distant sites^2,6,7^. Expression of p16INK4a (hereby referred to as p16) is the most commonly used marker of cellular senescence. p16 is not expressed in proliferating cells but induced by pro-senescence stimuli, marking transition to senescence and maintaining the senescence phenotype^2^.

While p16 expression is detected in essentially every aging tissue in mammals, recent studies demonstrate that cellular senescence in immune cells induces organismal senescence, onset of age-related diseases, and functional decline^8^. We have previously shown that p16 expression, measured in peripheral blood T lymphocytes of human study participants, is induced by a variety of age-promoting stimuli and correlates with frailty in cancer survivors^9,10^.

Soluble factors, secreted by senescent cells collectively known as the senescence-associated secretory phenotype (SASP), are thought to mediate pathophysiological effects of senescent cells^11^. While composition of the SASP varies greatly with both context (e.g., identity of stimuli, duration) and cell-type^12,13^, recent studies identified a subset of SASP proteins as a marker of health and clinical outcomes^14^.

CpG island methylation fingerprints have also recently been developed to measure aging^15–18^. Commonly referred to as epigenetic clocks of aging, they were developed to capture intrinsic (chronological age-based) as well as extrinsic (external damage that is highly variable between individuals) causes of aging. However, there is no consensus on which biological mechanisms they measure and each clock is composed of overlapping and distinct CpG loci and other features, that are proposed to capture biological aging processes.

While there is reported evidence that global epigenetic changes play a role in the establishment and maintenance of the senescence phenotype^19–21^, it is not known whether cellular senescence and signatures of methylation of the specific CpG islands that comprise various epigenetic clocks correlate in humans, *in vivo*. Here, in women with early-stage breast cancer and non-cancer controls, we performed a head-to-head comparison of the response of cellular senescence and five epigenetic clocks to cytotoxic chemotherapy, the most common and clinically-relevant intervention that accelerates biological aging and frailty in humans^9,10,22,23^. With an increase in interest in therapeutic interventions devised to slow aging, and, particularly, senolytic drugs that deplete senescent cells, it is imperative to understand how these biomarkers relate to each other.

## RESULTS

Participants diagnosed with early-stage breast cancer who were to undergo chemotherapy treatment were enrolled at UNC Hospitals. Forty-eight participants that had measurements of both p16 expression in peripheral blood T lymphocytes and methylation data before and 3-6 months after receiving chemotherapy were selected and analyzed (Figure 1 and Table 1). On average, these participants were 57 years (range 28-77). Seventy-seven percent of participants were white, 58% received doxorubicin-based chemotherapy with the rest receiving docetaxel and cyclophosphamide. The majority of participants (77%) received radiation therapy in addition to chemotherapy per standard of care.

**Figure 1.**
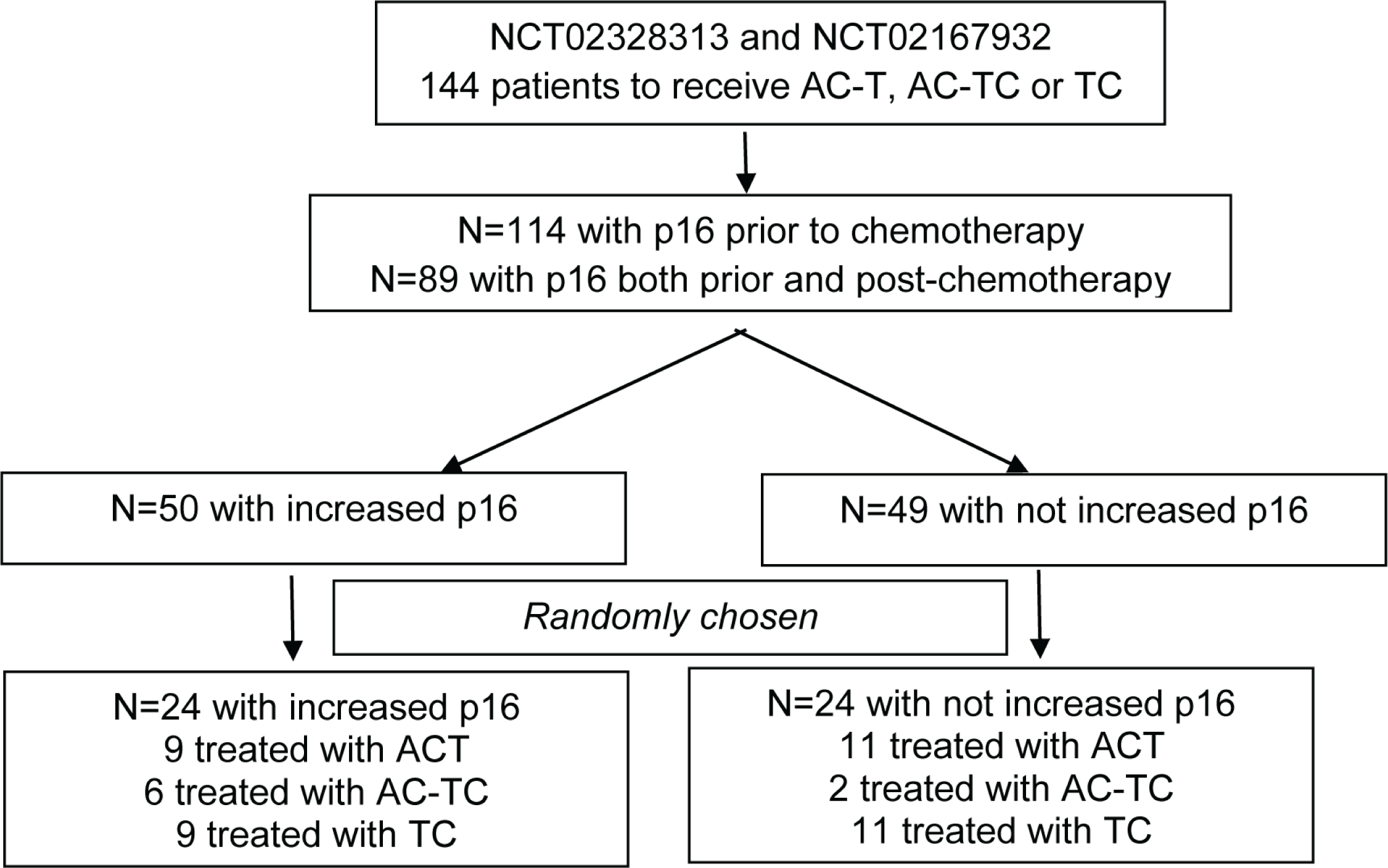
Consort flow diagram of the UNC cohort.

**Table 1:** Demographic and clinical characteristics of participants in the UNC Cohort.

| Variable | n=48 |
| --- | --- |
| **Demographics** |  |
| Age |  |
| Mean (SD) | 57 (13) |
| Range | 28 – 77 |
| BMI (kg/m<sup>2</sup>) |  |
| Mean (SD) | 28 (5.3) |
| Range | 20 - 38 |
| Comorbidities, n (%) |  |
| High blood pressure | 13 (27%) |
| Heart disease | 3 (6%) |
| Diabetes | 2 (4%) |
| Arthritis | 13 (27%) |
| Osteoporosis | 5 (10%) |
| Stomach or intestinal disorders | 12 (26%) |
| Race, n (%) |  |
| White | 37 (77%) |
| Black | 7 (15%) |
| Other | 4 (8%) |
| **Cancer characteristics** |  |
| Breast cancer stage, n (%) |  |
| I | 10 (21%) |
| II | 26 (54%) |
| III | 12 (25%) |
| Hormone Receptor status, n (%) |  |
| Positive | 30 (63%) |
| Negative | 18 (37%) |
| HER2, n (%) |  |
| Positive | 0 (0%) |
| Negative | 48 (100%) |
| Timing of chemotherapy, n (%) |  |
| Adjuvant | 29 (60%) |
| Neoadjuvant | 19 (40%) |
| Post-chemo Radiation, n (%) |  |
| No | 11 (23%) |
| Yes | 37 (77%) |
| Type of chemotherapy |  |
| AC-T (Doxorubicin/Cyclophosphamide plus Paclitaxel) | 20 (42%) |
| AC-TC (Doxorubicin/Cyclophosphamide plus Paclitaxel/Carboplatin) | 8 (17%) |
| TC (Docetaxel/Cyclophosphamide) | 20 (42%) |

### Chemotherapy induced an increase in p16 expression but little change in epigenetic clocks

Chemotherapy is a potent inducer of cellular senescence and is associated with accelerated functional decline and development of co-morbidities. When we compared expression of aging biomarkers before and three to six months after chemotherapy, p16 mRNA expression levels were increased by chemotherapy (p=0.003), but most epigenetic clocks were not, with the exception of DNAm-Hannum (p=0.02) (Figure 2). On average, p16 expression was increased by 0.7 in log2 units, the equivalent to 25 years of chronological aging^10^.

**Figure 2.**
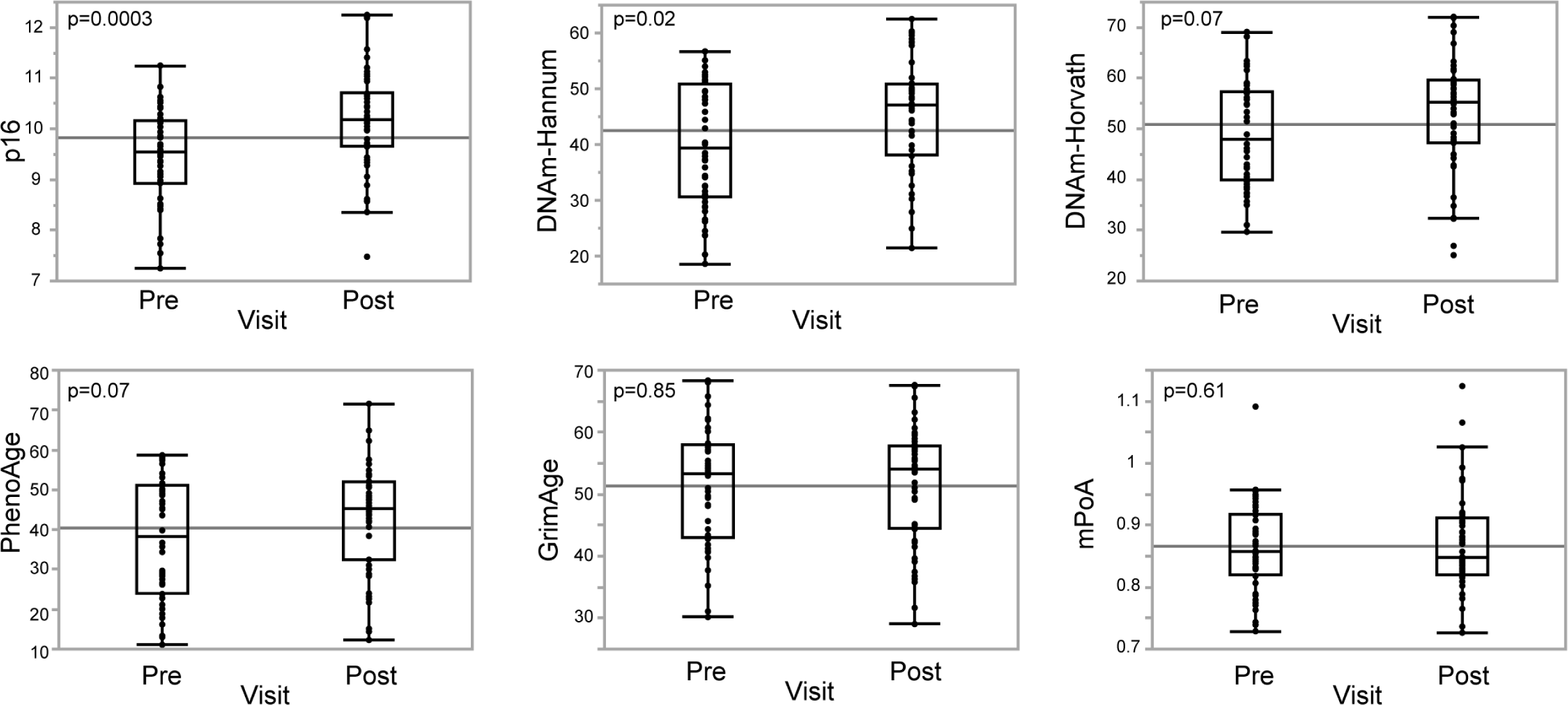
Chemotherapy-induced change in p16 and epigenetic clocks in participants in the UNC cohort.

To compare chemotherapy-induced changes in p16 and epigenetic clocks head-to-head, we stratified the cohort into participants whose p16 was induced by chemotherapy above the assay precision (n=24) or not (n=24). Then, we compared chemotherapy-induced changes in epigenetic clocks (differential between measures at follow-up and baseline) between the p16 groups. As shown in Figure 3, none of the epigenetic clocks changed with increased p16 expression.

**Figure 3.**
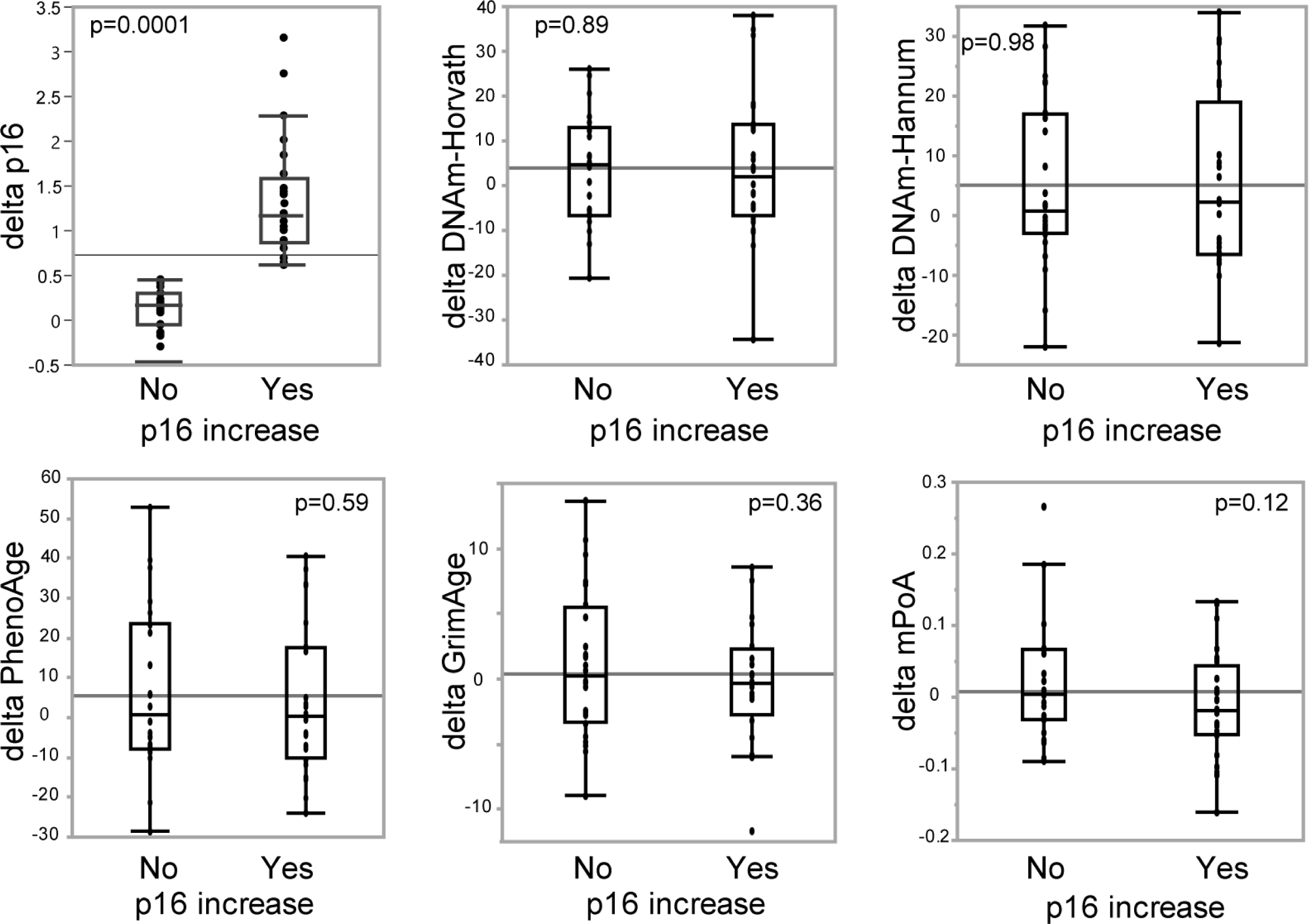
Correlation between chemotherapy-induced changes in epigenetic clocks and binary change in p16 in the UNC cohort.

### A subset of SASPs is induced with p16 by chemotherapy

To further describe chemotherapy-induced changes in senescence, we examined a panel of SASP proteins previously shown to be associated with morbidity and mortality^14^. As induction of individual components of SASP can be stimuli-specific, we limited analysis to samples from participants who received doxorubicin-based chemotherapy and had plasma samples available (n=20). Given the small sample size, we set statistical significance at p<0.1 (two-sided). Chemotherapy-induced changes in p16 correlated with increased levels of PARC, TNFRII, ICAM1, and TNFa (Table 2 and Figure 4). Interestingly, there was little to no association between baseline levels of p16 and baseline levels of these SASPs (Figure 5), suggesting that the association between p16 and SASP components is driven by chemotherapy-induced changes. Together, these data show that age-accelerating chemotherapy induces both a biomarker of cellular senescence, p16, and a subset of measured SASP components, but not epigenetic clocks.

**Figure 4.**
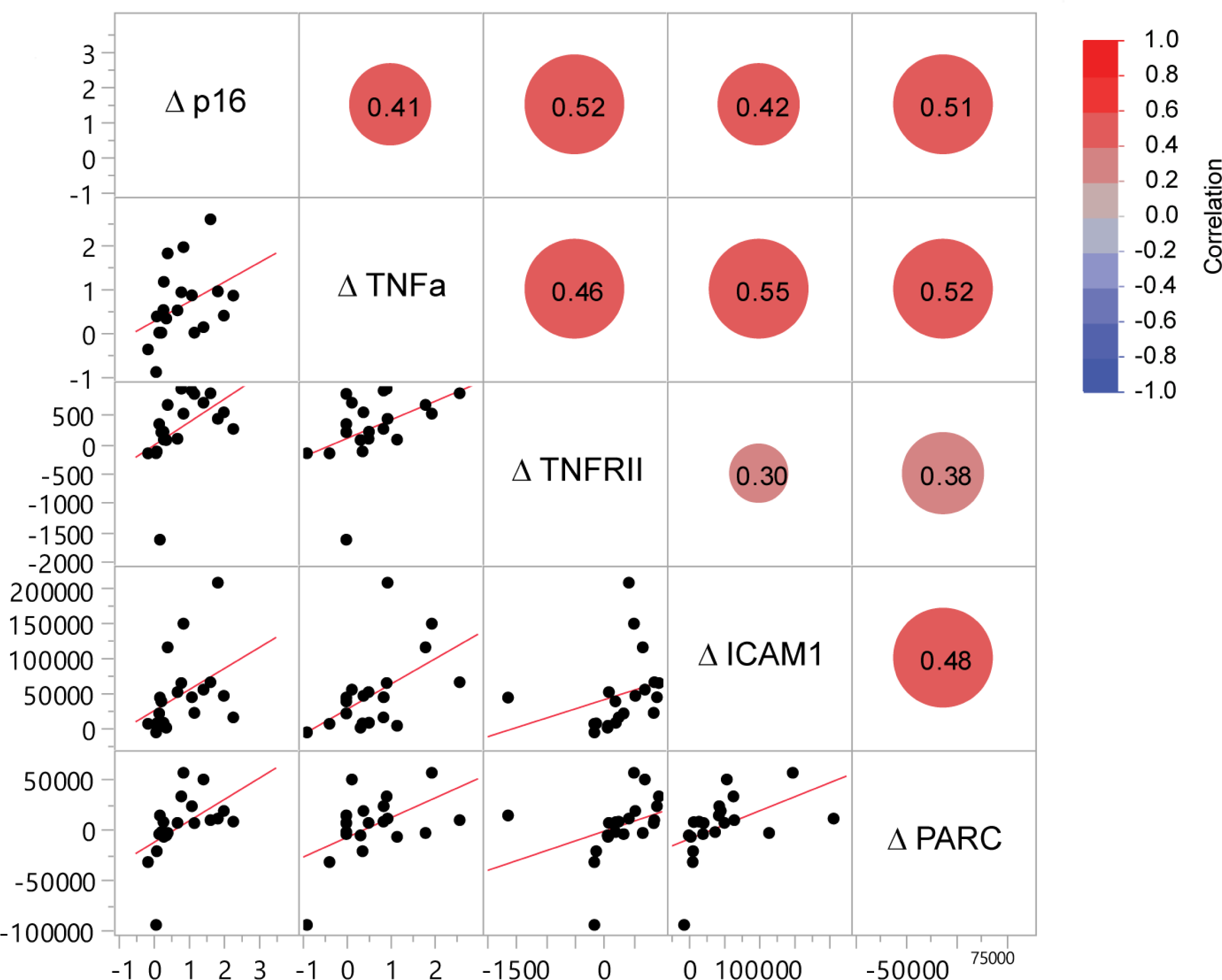
Correlation between chemotherapy-induced changes in p16 and SASPs in the UNC cohort.

**Figure 5.**
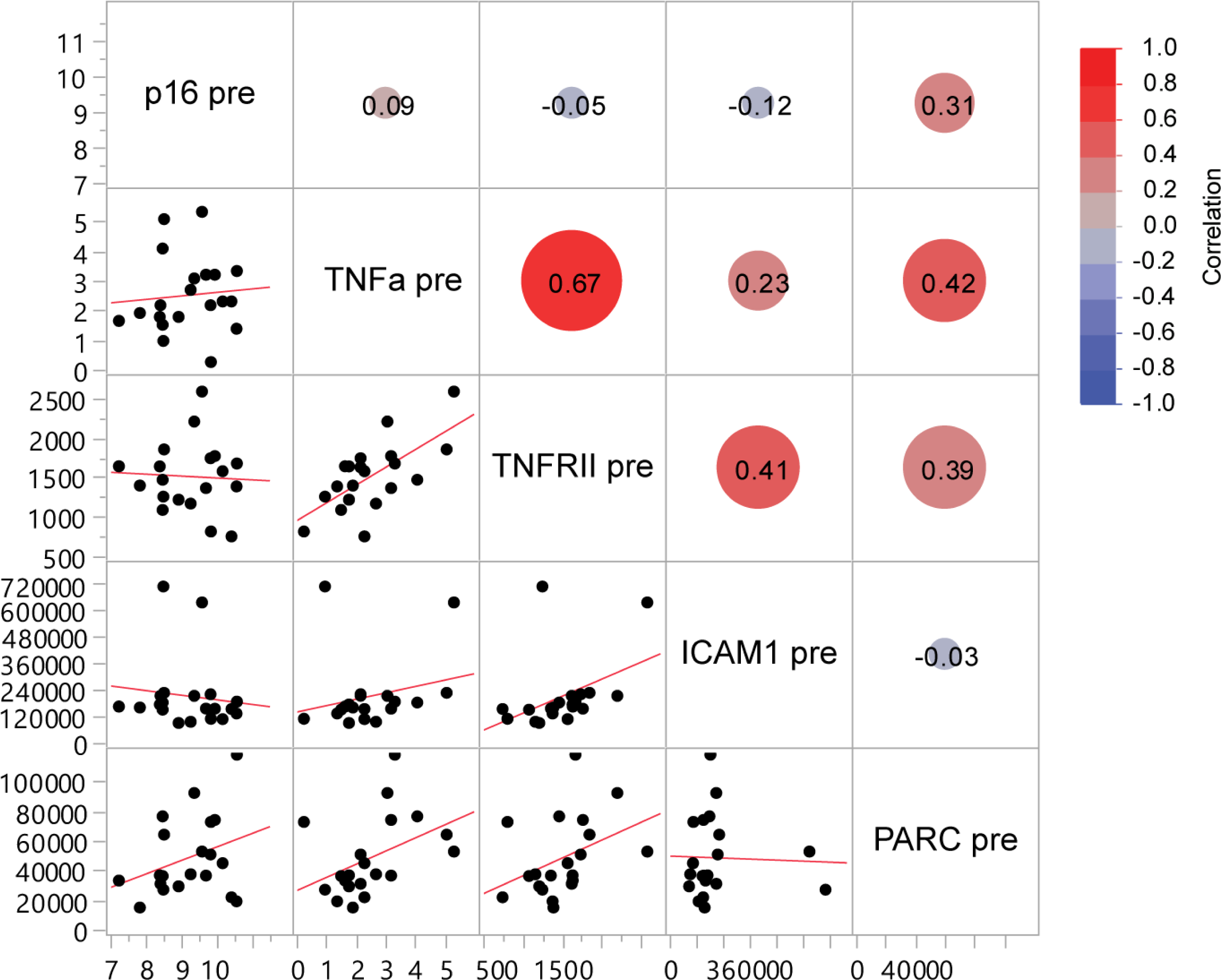
Correlation between p16 and SASPs prior to chemotherapy in the UNC cohort.

**Table 2.**
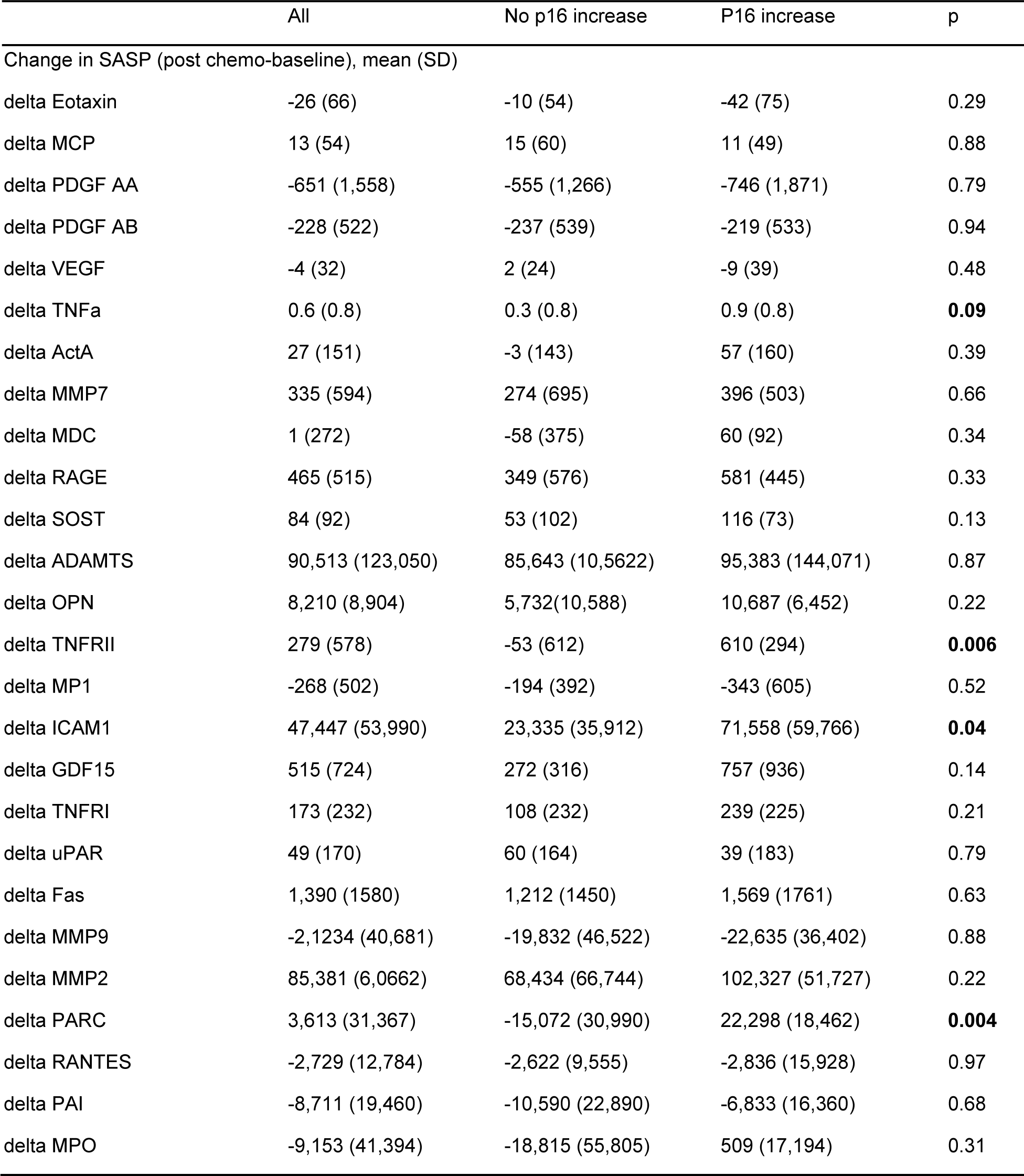
Correlation between chemotherapy-induced changes in SASPs and p16 in participants receiving doxorubicin-based chemotherapy in the UNC cohort.

### Correlation between p16 and DNA methylation clocks

While epigenetic clocks were not induced by chemotherapy, unlike the p16 senescence marker, we wanted to further examine the relationship between these markers prior to chemotherapy. The strongest association was detected between p16 expression and GrimAge. There was a weak association between p16 expression levels and clocks calibrated on chronological age to capture intrinsic aging (DNAm-Hannum r=0.22, p=0.02; and Horvath r=0.18, p=0.03). Clocks calibrated on chronological age and clinical characteristics to capture extrinsic aging were differentially associated with p16 expression with GrimAge being the strongest (r=0.31, p=0.0003), PhenoAge being weakly associated (r=0.14, p=0.03), and pace of aging (mPoA) not statistically significant (r=-0.01, p=0.8) (Figure 6). Non-surprisingly, there was a strong correlation between the chronological age clocks and among the three phenotypic clocks.

**Figure 6.**
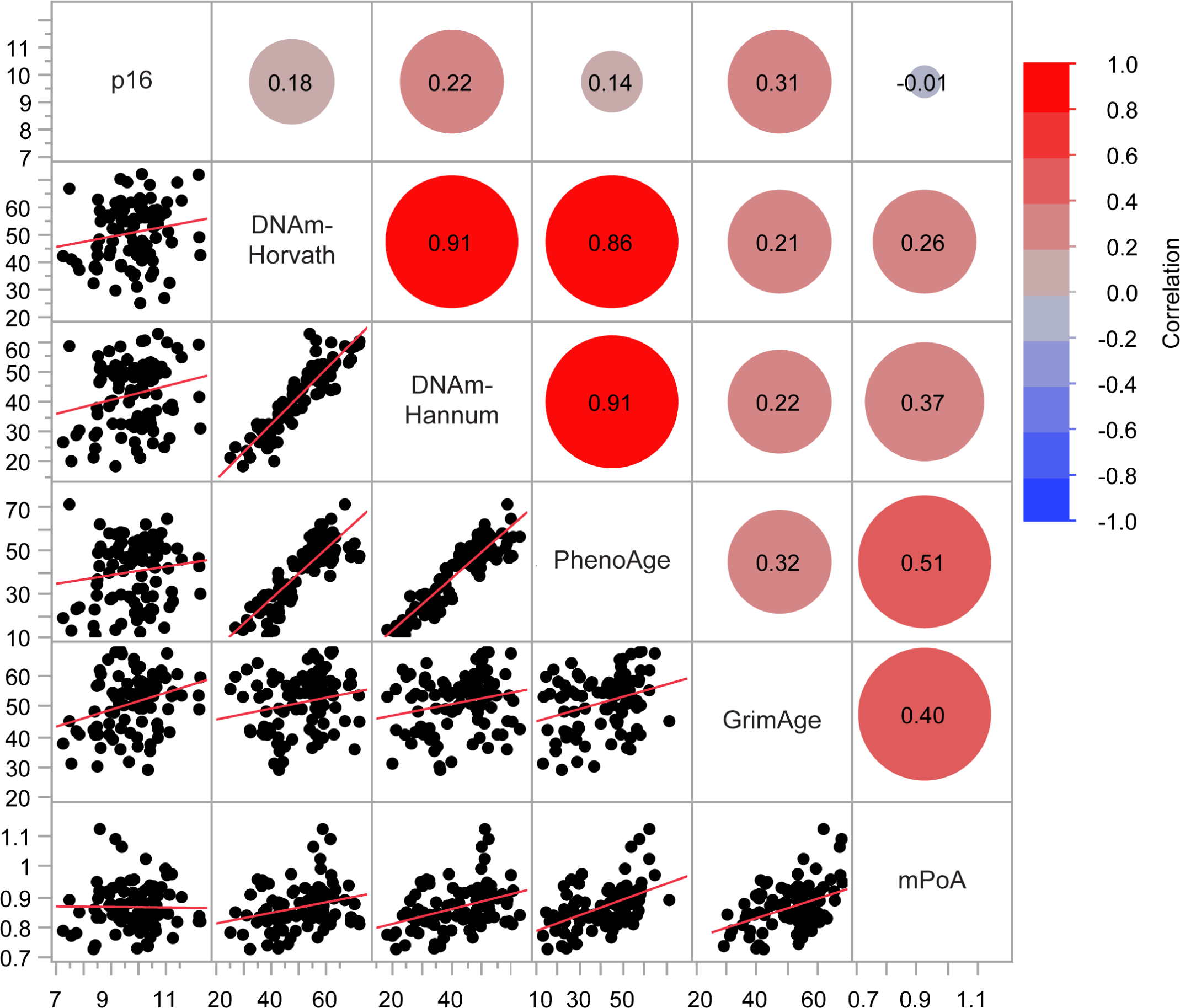
Scatterplot correlation matrix of p16 and epigenetic clocks prior to chemotherapy in the UNC cohort. The color of each correlation circle represents the correlation between each pair of variables on a scale from red (+1) to blue (-1). The size of each circle represents the significance test between the variables. A larger circle indicates a more significant relationship and the Pearson correlation coefficient is shown as a number.

Given the small cohort size used for the analysis of correlation between p16 and epigenetic clocks, we further extended our analyses to an independent cohort of participants recruited at the City of Hope. Table 3 summarizes key features of the City of Hope cohorts and the UNC cohort. First, we examined the correlation between p16 and epigenetic clocks (prior to any treatment for cancer) in a cohort of 251 participants diagnosed with early-stage breast cancer, similarly to the UNC cohort. All participants in the City of Hope cohort were 65 and older, and the median age was 71 years old (range 65-86) (Table 4), about 10 years older than the UNC cohort. Most participants were non-Hispanic white (82.1%) or black (11.2%). As in the UNC cohort, there was a weak association between p16 expression and DNAm-Hannum (r=0.15, p=0.02) and stronger association with PhenoAge and GrimAge (r=0.2, p=0.002 and r=0.13, p=0.04, respectively) (Figure 7). p16 was not associated with DNAm-Horvath or mPoA.

**Figure 7.**
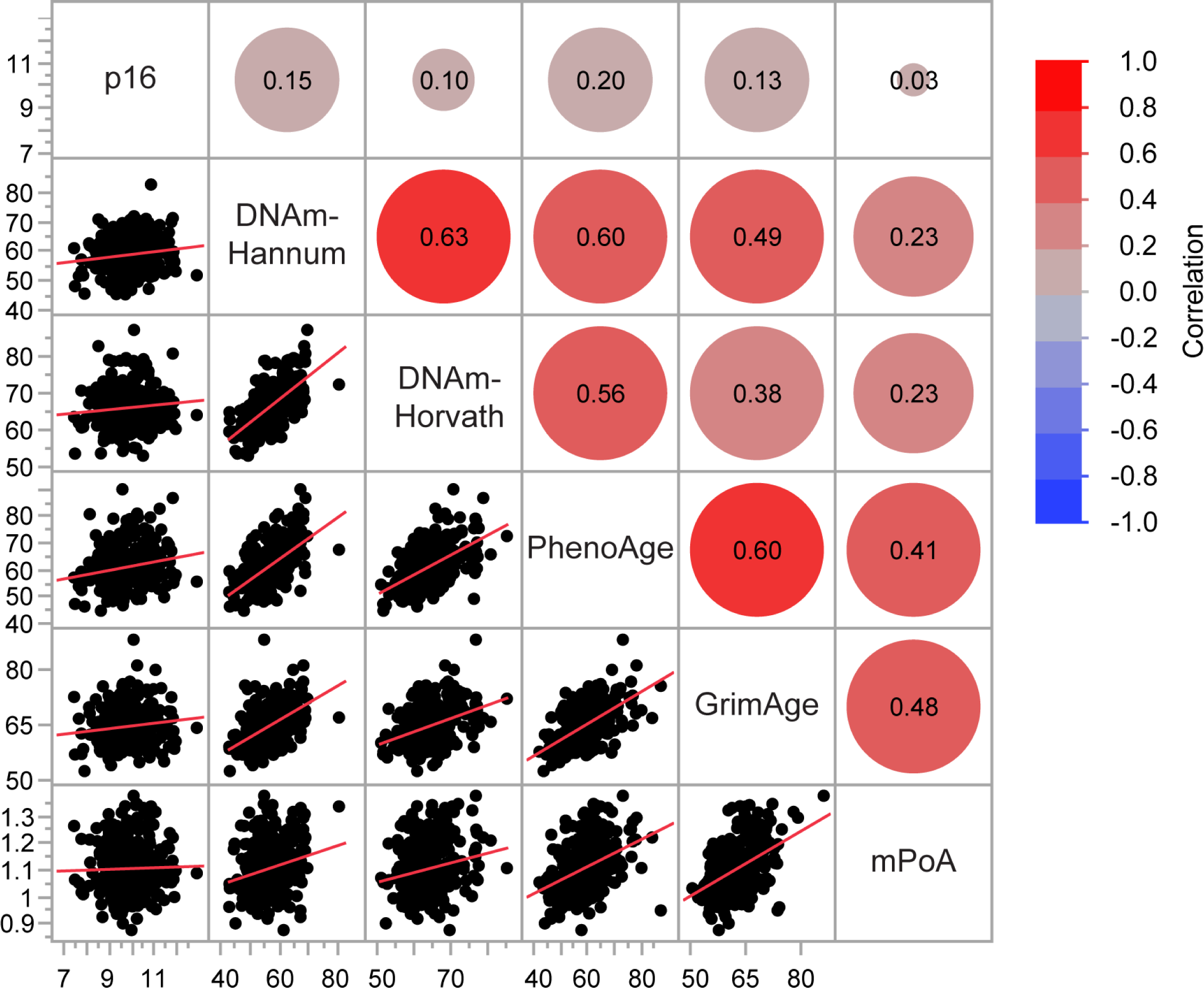
Scatterplot correlation matrix of p16 and epigenetic clocks in participants with cancer prior to receiving cancer treatment. The color of each correlation circle represents the correlation between each pair of variables on a scale from red (+1) to blue (-1). The size of each circle represents the significance test between the variables. A larger circle indicates a more significant relationship and the Pearson correlation coefficient is shown as a number.

**Table 3.** Overview of main characteristics of cohorts used in the analysis.

|  | UNC Cohort | COH cohort |  |
| --- | --- | --- | --- |
| No. participants | 48 | 251 | 49 |
| Study site | UNC Hospitals | City of Hope | City of Hope |
| Participants | early-stage breast cancer diagnosis, ages 28-77 | early-stage breast cancer diagnosis, ages 65-86 | age-matched volunteers without cancer, ages 65-90 |
| Aging biomarkers measured | p16 cellular senescence<br>Epigenetic clocks<br>SASPs | p16 cellular senescence<br>Epigenetic clocks | p16 cellular senescence<br>Epigenetic clocks |
| Time of measurement | Baseline and post-chemotherapy | Baseline only | Baseline only |

**Table 4.** Characteristics of participants diagnosed with cancer in the City of Hope cohort.

|  | Breast Cancer<br>(N=251) |
| --- | --- |
| **Age** |  |
| Mean (SD) | 71.2 (4.7) |
| Range | 65-86 |
| **BMI** |  |
| Mean (SD) | 29.5 (5.8) |
| Range | 17.9-51.2 |
| **Gender** |  |
| Female, n (%) | 251 (100%) |
| **Race/Ethnicity, n (%)** |  |
| Non-Hispanic white | 206 (82.1%) |
| Black | 28 (11.2%) |
| Asian | 5 (2.0%) |
| Hispanic | 11 (4.4%) |
| Other | 1 (0.4%) |
| **Comorbidities, n (%)** |  |
| High blood pressure | 148 (59) |
| Heart disease | 34 (13.5) |
| Diabetes | 47 (18.7) |
| Arthritis | 128 (51.0) |
| Osteoporosis | 50 (19.9) |
| Stomach or intestinal disorders | 48 (19.1) |
| **Prior cancer surgery, n (%)** | 207 (82.5%) |
| **Cancer Stage, n (%)** |  |
| I | 100 (39.8%) |
| II | 107 (42.6%) |
| III | 44 (17.5%) |
| **Tumor, n (%)** |  |
| HR+/HER2+ | 35 (13.9%) |
| HR+/HER2- | 145 (57.8%) |
| HR-/HER2+ | 22 (8.8%) |
| Triple negative | 49 (19.5%) |

Next, we examined the association between p16 and epigenetic clocks in a cohort of volunteers that did not have history of cancer and were age-matched to the City of Hope cancer cohort (Table 5). Again, similarly to the UNC cohort, there was a weak association between p16 and intrinsic aging clocks (Horvath r=0.36, p=0.04, Hannum r=0.3, p=0.01) and extrinsic aging clocks PhenoAge and GrimAge (r=0.36, p=0.01 and r=0.33, p=0.02, respectively), but not mPoA (Figure 8).

**Figure 8.**
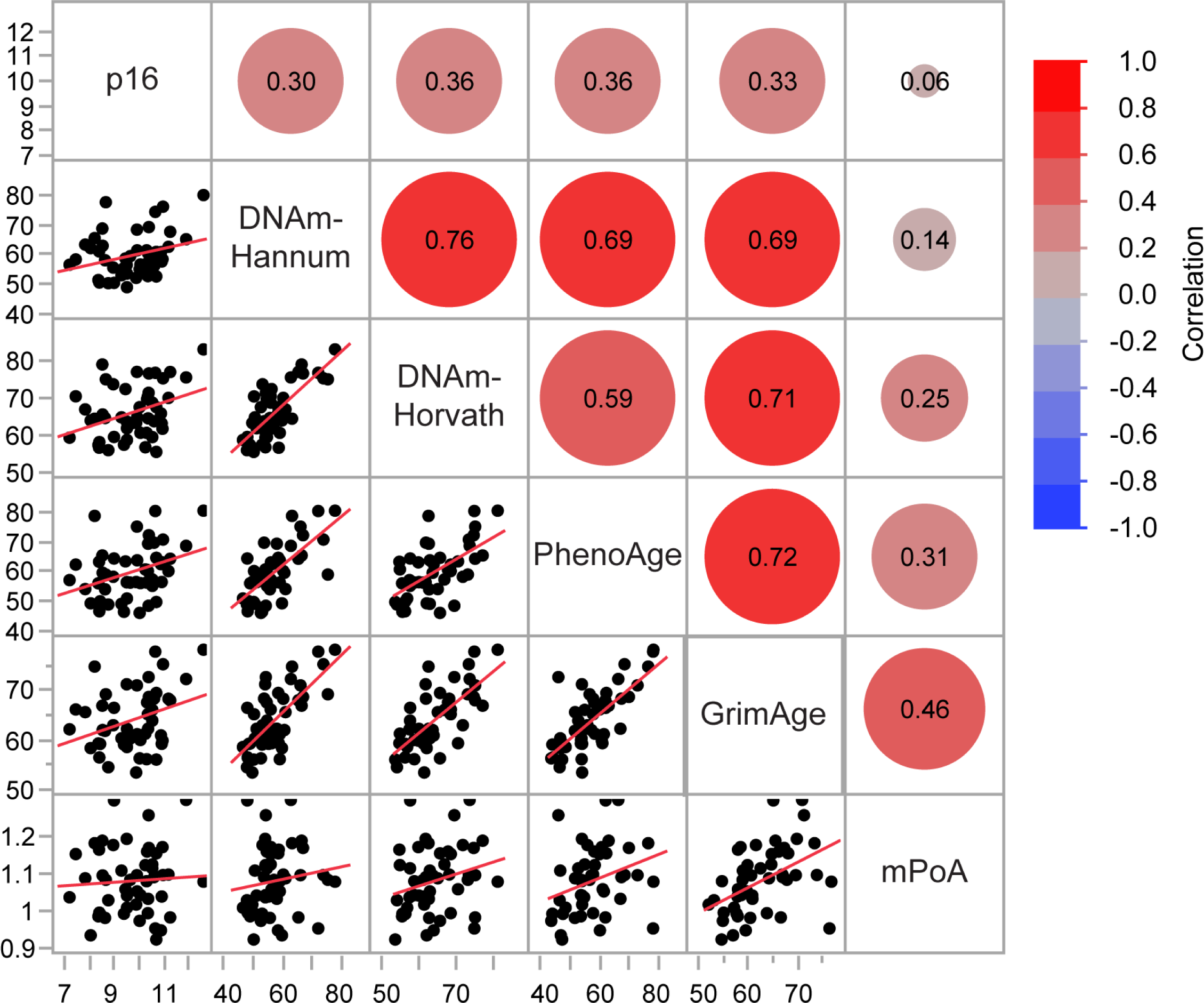
Scatterplot correlation matrix of p16 and epigenetic clocks in participants with no history of cancer. The color of each correlation circle represents the correlation between each pair of variables on a scale from red (+1) to blue (-1). The size of each circle represents the significance test between the variables. A larger circle indicates a more significant relationship and the Pearson correlation coefficient is shown as a number.

**Table 5.** Characteristics of participants without cancer in the City of Hope cohort.

|  | No Cancer<br>(N=49) |
| --- | --- |
| **Age** |  |
| Mean (SD) | 72.2 (6.10) |
| Range | 65-90 |
| **BMI** |  |
| Mean (SD) | 28.1 (6.1) |
| Range | 18.4-41.7 |
| **Gender** |  |
| Female, n (%) | 49 (100%) |
| **Race/Ethnicity, n (%)** |  |
| Non-Hispanic white | 33 (67.4%) |
| Black | 9 (18.4%) |
| Asian | 1 (2.0%) |
| Hispanic | 6 (12.2%) |
| Other | 0 (0%) |
| **Comorbidities, n (%)** |  |
| High blood pressure | 24 (49) |
| Heart disease | 3 (6.1) |
| Diabetes | 6 (12.2) |
| Arthritis | 29 (59.2) |
| Osteoporosis | 10 (20.4) |
| Stomach or intestinal disorders | 9 (18.4) |

A recent study found that aging may be accelerated, when assessed through epigenetic clocks, in people with some cancers compared to cancer-free study participants^24^. To determine if participants with the early-stage breast cancer have distinct expression of cellular senescence biomarker and epigenetic clocks, we plotted p16 or epigenetic clocks as a function of chronologic age and compared regression lines (Figure 9). Regression lines in participants with and without cancer were not significantly different for p16, DNAm-Hannum, DNAm-Horvath and mPoA. Epigenetic ages in GrimAge and PhenoAge algorithms were statistically higher in participants with cancer than without cancer (p=0.02 and 0.03, respectively). Since epigenetic clocks can be influenced by race, ethnicity, and obesity/BMI^15,25^, adjusted values for each of epigenetic clock were used for comparison between subgroups as well. Difference in PhenoAge and GrimAge between cancer and no-cancer groups remained statistically significant when adjusted for race, ethnicity, and BMI (p=0.04 and p=0.01, respectively; data not shown).

**Figure 9:** Regression analysis of p16 expression levels and epigenetic clocks vs chronological age in participants with or without cancer history in the City of Hope cohort.

## DISCUSSION

We found that the cellular senescence marker p16 but not epigenetic clocks of aging responded to a clinically relevant inducer of human aging, cytotoxic chemotherapy. We also found weak to no correlation between cellular senescence and five of the most commonly used epigenetic clocks in patients prior to chemotherapy or in an age-matched cohort of donors without history of cancer, suggesting that there is a general discordance between measures of cellular senescence and epigenetic clocks. GrimAge and PhenoAge had the strongest association with p16 with a correlation coefficient of <0.3.

There are two general areas of consideration in order to contextualize our findings. First, cellular senescence, but not epigenetic clocks increased by age-accelerating chemotherapy treatments. Our observations appear counter to the findings of Qin et al where an increase in PhenoAge was detected in survivors of childhood cancers^26^. However, epigenetic clock measurements in the study of Qin et al were taken about two decades after the end of treatment. Therefore, it is possible that epigenetic clocks do change in response to pro-aging effects of chemotherapy but the effect is indirect and is not seen within the 3-6 months follow-up window of this study.

Second, existing epigenetic aging clocks may not capture changes in the epigenome that characterize cancer risk and chemotherapy-induced age acceleration. We found no difference in any of the epigenetic clocks in patients with early-stage breast cancer and age-matched donors consistent with the findings of Berstein et al that epigenetic clocks cannot identify people at risk for a wide range of cancers^24^. The lack of upregulation of epigenetic clocks shortly after chemotherapy is more likely to be an indication of the inability of aging clocks to capture relevant epigenetic changes, rather than the lack of epigenetic changes altogether. In fact, Yao and colleagues found that chemotherapy induces early and substantial changes to leukocyte DNA methylation as well as proportions of immune cells, including B cells and CD4+ T cells in early-stage breast cancer patients^27^. Analysis of epigenetic clocks is also confounded by changes in immune cell subtypes, as elegantly demonstrated by Tomusiak et al^28^, and therefore, chemotherapy-induced perturbations of the immune system could make measures of commonly used clocks unreliable.

Overall, identification of biomarkers of aging that can be used to characterize aging phenotypes or “hallmarks” is a rapidly expanding field. Understanding mechanisms involved in aging biomarker regulation as well as demonstrating their causative relationship to disease initiation, progression, and aging-related functional decline is imperative. Biomarkers that are clearly induced in clinical models of human aging are required to identify and measure the efficacy of clinical interventions, including novel pharmacotherapies that may be either pro- or anti-aging.

## MATERIALS AND METHODS

### Study Participants and Treatments

To examine the effect of chemotherapy treatment on expression level of biomarkers of aging, we used samples collected from patients 18 years or older enrolled into two studies of patients with early-stage breast cancer and conducted at the University of North Carolina: NCT02328313 and NCT02167932. Standard practice regimens were chosen by participants and their oncologists based on cancer stage and phenotype: regimens containing doxorubicin in combination with cyclophosphamide, paclitaxel or carboplatin (AC-T or AC-TC), or regimens containing docetaxel and cyclophosphamide (TC). Patients who had p16 measurements both before an after chemotherapy were considered (see Figure 2) and a subset of 48 patients was chosen randomly for further analysis of methylation. For analysis comparing changes in p16 and SASPs, 20 paired pre- and post-chemotherapy plasma samples from participants who received doxorubicin-based chemotherapy (AC-T) were used.

Furthermore, we used data from The “Clinical and Biological Predictors of Chemotherapy Toxicity in Older Adults” study which was led by the City of Hope (City of Hope cohort) and that accrued patients from 16 participating U.S. institutions (NCT01472094). Three groups of patients were recruited for this study: 1) chemotherapy patients, aged ≥65, who had a diagnosis of stage I-III breast cancer and were scheduled to receive adjuvant/neoadjuvant chemotherapy (n=501); 2) breast cancer patients who were not planning to receive adjuvant or neo-adjuvant chemotherapy and age-matched to the chemotherapy group with the same disease eligibility criteria (n=100); 3) participants without history of cancer as an age-matched no-cancer cohort (n=100). All cancer patients received treatment per provider discretion. Groups 1 and 2 were grouped together as a “diagnosed with cancer” group. Among 701 participants acoss the entire City of Hope cohort, 319 had both p16 and epigenetic clock data measured prior to chemotherapy or other cancer treatment or at enrollment (for a no-cancer cohort) were included in this analysis. 19 patients were excluded from analyses in the cancer group as they had chemotherapy or radiation prior to participation in this study and p16 sample collection. The resulting cohort of 251 participants diagnosed with cancer and 49 participants without cancer were used for analyses in this manuscript.

All these studies were approved by the Protocol Review Committee as well as institutional review boards at all study sites, with written informed consent given by all participants.

### Measures of biomarkers of aging

### p16 expression levels

Whole peripheral blood was drawn into EDTA tubes prior to and at the end of chemotherapy treatment (three months to six months from the end of treatment). T cells were isolated using negative selection (RosetteSep, StemCell Technologies) and analyzed for p16 gene expression using TaqMan probes and real-time quantitative PCR of reverse-transcribed cDNA, essentially as described previously^9,10^. Positive and negative controls were included in each run to monitor assay performance; overall precision of p16 measurement was 0.8Ct.

### Epigenetic clocks

*University of North Carolina Cohort*- For methylation analysis, we chose 48 participants randomly (see Figure 1) where p16 was increased by chemotherapy in 24 participants and not increased in the other 24. 96 samples (paired pre- and post-chemotherapy samples) were then used in methylation analysis. A sample size of 96 was used to avoid potential batch variation. Genomic DNA was isolated from whole blood samples stored at −80° C using commercially avaliable kit (Zymo Technologies). DNA concentration was determined using Nanodrop 8000 and samples were prepared for analysis in a single 96-well plate at the UCLA Neurosciences Genomics Core per their instructions. Raw methylation image files were processed using the *minfi* and EnMix packages^29,30^ in R. Normalized values were used to calculate epigenetic ages using the online epigenetic age calculator (https://dnamage.genetics.ucla.edu/new)^15^. DunedinPoAm38 R package was used to calculate DunedinPoAm38 DNAm age^18^ (mPoA).

*City of Hope cohort*- Genomic DNA was isolated from whole blood samples stored at −80° C. DNA concentration was determined using Nanodrop 8000 and samples were prepared for analysis in 96-well plates. To measure methylation, 500 ng DNA from each sample was assayed using the EPIC Bead Chip (Illumina Inc., CA, USA) in the City of Hope Integrative Genomics Core. Raw methylation image files were processed using the *minfi* and EnMix packages ^29,30^ in R. The “preprocessNoob” method was used for normalization^31^. The type I and type II probe bias was adjusted using the method of regression on correlated probes^32^. Batch effects related to laboratory technical variation were adjusted using the top three surrogate variables, accounting for 96% of array control-probe variation, estimated based on the singular value decomposition (SVD) method^29^. Batch adjusted residuals were used for DNAm age calculation using the online epigenetic age calculator which includes the four methylation clocks analyzed (https://dnamage.genetics.ucla.edu/new)^15^. DunedinPoAm38 R package was used to calculate DunedinPoAm38 DNAm age^18^ (mPoA).

### SASP expression

Plasma samples that were isolated from whole blood by centrifuging on the Ficoll gradient and stored at −80° C were used to measure expression of SASPs. Concentrations of target proteins in patient plasma samples (250 ul) were quantified using commercially available multiplex magnetic bead immunoassays (R&D Systems) based on Luminex xMAP multianalyte profiling platform and analyzed on a MAGPIX System (Merck Millipore) and, for activin A, an enzyme-linked immunoabsorbent assay (R&D Systems). All assays were performed according to the manufacturer’s protocols^14,33^.

### Statistical Analysis

*D*escriptive statistics were used to summarize demographic, disease and clinical characteristics for each study group. Linear regression and pearson correlation analyses were used to establlish correlation between p16 and epigenetic clocks or SASP expression levels. Student’s t-test was used to compare change in methylation clocks with chage in p16 as a binary variable. p value of less than 0.05 was considered to be significant unless noted otherwise.

## AUTHOR CONTRIBUTIONS

**Conception and design:** Muss, Mitin

**Provision of study materials or participants:** Muss, Nyrop, Sedrak

**Collection and assembly of data:** Knecht, Strum, Nyrop, Mitin, LeBrasseur, Sun, White

**Data analysis and interpretation:** Mitin, LeBrasseur, Muss, Sun, Ding, Neuhausen, Sedrak

**Manuscript writing, review, and editing:** All authors

## Data Availability

Data can be requested from the senior author

## ACKNOWLEDGEMENTS

We would like to thank Dr Terrie Moffitt for discussions and review of the manuscript.

## CONFLICTS OF INTEREST

NM is a co-founder of Sapere Bio. NM, SLS, and AK are equity holders in the company and inventors on intellectual property applications. Mayo Clinic and NKL have relevant technology licensed to a commercial entity. This research has been reviewed by the Mayo Clinic Conflict of Interest Review Board and is being conducted in compliance with Mayo Clinic Conflict of Interest policies. Other authors report no conflict of interest.

## FUNDING

This work was supported by grants from Breast Cancer Research Foundation, New York, NY, the Kay Yow Cancer Fund, Cary, NC, the UNC Lineberger Comprehensive Cancer Center/University Cancer Research Fund and NIH/NCI R01 CA203023 to HM. NKL is supported by the NIH/NIA, including R01 AG55529 and R56 AG60907, and the Glenn Foundation for Medical Research. The funding sources had no involvement in the study design or in the collection, analysis and interpretation of data. The funding sources also had no involvement in the writing of this report or in the decision to submit the report for publication.

## Notes

### Author Declarations

Two Trials used in this study, NCT#02328313 and 02167932 that included the University of North Carolina Cohort were approved but the Institutional Review Board of the University of North Carolina, Chapel Hill, NC The City of Hope Cohort included patients from NCT# 0142094 and was approved by the Institutional Review Boards of: City of Hope Medical Center, Duarte, California Yale University New Haven, Connecticut University of Chicago, Chicago, Illinois, Massachusetts General Hospital, Boston, Massachusetts Dana-Farber Cancer Institute, Boston, Massachusetts Washington University St. Louis, Missouri Roswell Park Cancer Institute, Buffalo, New York Hofstra-North-Long Island Jewish Cancer Institute, New Hyde Park, New York Memorial Sloan-Kettering Cancer Center, New York, New York University of Rochester Medical Center, Rochester, New York. University of North Carolina, Chapel Hill, North Carolina Wake Forest University, Winston-Salem, North Carolina Case Western Reserve University, Cleveland, Ohio Thomas Jefferson University, Philadelphia, Pennsylvania Fox Chase Cancer Center, Philadelphia, Pennsylvania Rhode Island Hospital, Providence, Rhode Island University of Virginia Health System, Charlottesville, Virginia

